# A Genome-Wide Association Study of the Frailty Index Highlights Synaptic Pathways in Aging

**DOI:** 10.1101/19007559

**Authors:** Janice L Atkins, Juulia Jylhävä, Nancy L Pedersen, Patrik K Magnusson, Yi Lu, Yunzhang Wang, Sara Hägg, David Melzer, Dylan M Williams, Luke C Pilling

**Affiliations:** Epidemiology and Public Health Group, University of Exeter Medical School, RILD Building, Barrack Road, Exeter, UK; Department of Medical Epidemiology and Biostatistics, Karolinska Institutet, Stockholm, Sweden; Department of Psychology, University of Southern California, Los Angeles, CA, USA; Center on Aging, University of Connecticut, 263 Farmington Avenue, Farmington, CT, USA; MRC Unit for Lifelong Health and Ageing at UCL, University College London, London, UK

**Keywords:** Aging, Frailty, Frailty Index, Genetics, UK Biobank

## Abstract

Frailty is a common geriatric syndrome, strongly associated with disability, mortality and hospitalisation. The mechanisms underlying frailty are multifactorial and not well understood, but a genetic basis has been suggested with heritability estimates between 19 and 45%. Understanding the genetic determinants and biological mechanisms underpinning frailty may help to delay or even prevent frailty. We performed a genome-wide association study (GWAS) of a frailty index (FI) in European descent participants from UK Biobank (*n*=164,610, aged 60-70 years). FI calculation was based on 49 self-reported items on symptoms, disabilities and diagnosed diseases. We identified 26 independent genetic signals at 24 loci associated with the FI (*p*<5*10^−8^). Many of these loci have previously been associated with traits such as body mass index, cardiovascular disease, smoking, HLA proteins, depression and neuroticism; however, three appear to be novel. The estimated single nucleotide polymorphism (SNP) heritability of the FI was 14% (0.14, SE 0.006). A genetic risk score for the FI, derived solely from the UK Biobank data, was significantly associated with FI in the Swedish TwinGene study (*n*=10,616, beta: 0.11, 95% CI: 0.02-0.20, *p*=0.015). In pathway analysis, genes associated with synapse function were significantly enriched (*p*<3*10^−6^). We also used Mendelian randomization to identify modifiable traits and exposures that may affect the risk of frailty, with a higher educational attainment genetic risk score being associated with a lower risk of frailty. Risk of frailty is influenced by many genetic factors, including well-known disease risk factors and mental health, with particular emphasis on synapse maintenance pathways.

## INTRODUCTION

Frailty is a common geriatric syndrome involving multi-system impairment which is associated with increased vulnerability to stressors (Dodds & Sayer 2016). Frailty is a major public health issue in geriatric populations and is becoming increasingly common with aging demography (Morley 2016). Prevalence is estimated as 17.4% worldwide in community-dwelling older adults aged 60 and above (Siriwardhana et al. 2018). Many frailty definitions exist but two of the most commonly used are the frailty phenotype (FP) (Fried et al. 2001) and the frailty index (FI) (Mitnitski et al. 2001). The FP is a clinical syndrome based on the presence of three of five physical components (exhaustion, weakness, slow walking speed, unintentional weight loss and low physical activity), whereas the FI is based on the accumulation of a number of health deficits during the life course. While both measures are related to adverse aging outcomes, the FI is better at discriminating at the lower to middle end of the frailty continuum, making it better suited for measuring frailty in seniors at younger age (Blodgett et al. 2015).

Frailty is associated with many negative health outcomes including disability, mobility limitations, a variety of chronic diseases and hospitalisation, and it is a strong predictor of mortality (Vermeiren et al. 2016) (Kojima et al. 2018) (Chang & Lin 2015). The mechanisms underlying frailty are likely multifactorial but not well understood. Studies have suggested a genetic basis, with heritability estimates between 19 and 45% (Livshits, Lochlainn, et al. 2018) (Murabito et al. 2012) (Young et al. 2016). Candidate gene association studies for frailty have suggested the involvement of genes in inflammatory pathways (Viña et al. 2016), including tumour necrosis factor (Mekli et al. 2016) and interleukin-18 (Mekli et al. 2015). Large-scale GWAS are needed to better understand the genetic determinants of frailty and associated biological pathways. These might provide insights for interventions to prevent and delay frailty and hence promote healthier and more independent aging.

We undertook a GWAS of frailty, measured using the FI, in 60 to 70 year old participants of European descent from the UK Biobank cohort, with replication in the Swedish TwinGene cohort study. To gain insights into the functional relevance of emergent genetic associations, we examined the relationship of the frailty risk loci with circulating proteins and tissue-specific gene expression and epigenetic profiles. We also leveraged the GWAS findings in Mendelian randomization analyses to identify modifiable physiological, lifestyle or environmental traits that could be targeted by clinical and/or public health interventions to mitigate frailty risk.

## RESULTS

### Study characteristics

In this GWAS, we included 164,610 UK Biobank participants of European descent aged 60 to 70 years at baseline (mean 64.1, SD 2.8), which included 84,819 females (51.3%). The FI score ranged from zero to 27, from 49 deficits in total, and the mean proportion of deficits was 0.129 (SD 0.075), with a slightly higher mean score in females (0.132, SD 0.076) compared to males (0.125, SD 0.074).

The TwinGene participants (*n*=10,616) – whose data were used in replication testing of GWAS findings from UK Biobank – were Swedish nationals aged 41 to 87 years (mean 58.3, SD 7.9) with 5,577 females (52.5%), and all of European descent. The TwinGene FI score ranged from zero to 26.25 deficits (44 considered in total), and the mean proportion of deficits was 0.121 (SD 0.080). The Swedish Adoption/Twin Study of Aging (SATSA) participants (*n*=368) – whose data were used in DNA methylation-related follow-up analyses – were all of European descent, aged 48 to 93 years (mean 68.6, SD 9.6), with 223 (60.6%) females. The SATSA FI score ranged from zero to 19 deficits (42 considered in total), and the mean proportion of deficits was 0.100 (SD 0.087).

### GWAS of the FI

We identified 26 independent signals associated (*p*<5*10^−8^) with the FI (*n*= 5,849 associated variants in total), at 24 loci (see Table 1 and Figure 1). Eleven variants were associated at significance levels substantially below a multiple testing-corrected *p*-value threshold of 3*10^− 9^ (0.05 corrected for 16.4 million autosomal variants included in the analysis), although this is likely to be too conservative given the correlated nature of genetic variants. The Lambda GC (genomic control) value was high (1.37; see Supplementary Figure 1 for QQ plot), however the intercept from Linkage Disequilibrium Score Regression (LDSR) analysis was 1.02 (SE 0.009), indicating that the inflation in test statistics is mainly due to polygenicity (many variants with small effects on frailty), and the bias potentially due to population stratification is minimal (Bulik-Sullivan et al. 2015). The single nucleotide polymorphism (SNP)-based heritability (*h*^*2*^_*g*_) of the FI was estimated to be 0.14 (SE 0.006), i.e. 14%, by LDSR. Full GWAS summary statistics are available to download (https://dx.doi.org/10.6084/m9.figshare.9204998).

**Table 1.**
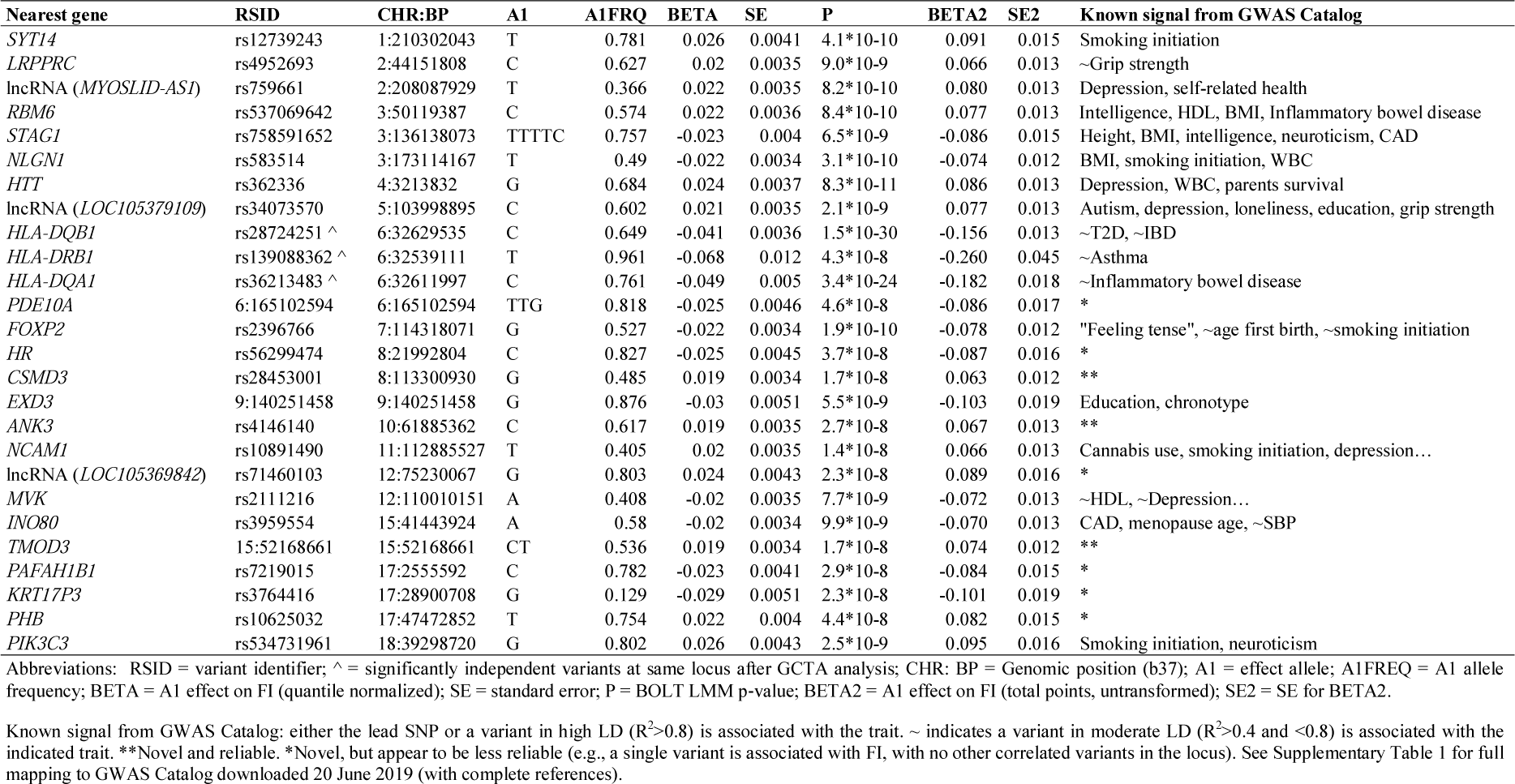
GWAS associations with Frailty Index in UK Biobank

| Nearest gene | RSID | CHR:BP | A1 | A1FRQ | BETA | SE | P | BETA2 | SE2 | Known signal from GWAS Catalog |
| --- | --- | --- | --- | --- | --- | --- | --- | --- | --- | --- |
| *SYT14* | rs12739243 | 1:210302043 | T | 0.781 | 0.026 | 0.0041 | 4.1*10<sup>-10</sup> | 0.091 | 0.015 | Smoking initiation |
| *LRPPRC* | rs4952693 | 2:44151808 | C | 0.627 | 0.02 | 0.0035 | 9.0*10<sup>-9</sup> | 0.066 | 0.013 | ~Grip strength |
| lncRNA (*MYOSLID-AS1*) | rs759661 | 2:208087929 | T | 0.366 | 0.022 | 0.0035 | 8.2*10<sup>-10</sup> | 0.080 | 0.013 | Depression, self-related health |
| *RBM6* | rs537069642 | 3:50119387 | C | 0.574 | 0.022 | 0.0036 | 8.4*10<sup>-10</sup> | 0.077 | 0.013 | Intelligence, HDL, BMI, Inflammatory bowel disease |
| *STAG1* | rs758591652 | 3:136138073 | TTTTC | 0.757 | -0.023 | 0.004 | 6.5*10<sup>-9</sup> | -0.086 | 0.015 | Height, BMI, intelligence, neuroticism, CAD |
| *NLGN1* | rs583514 | 3:173114167 | T | 0.49 | -0.022 | 0.0034 | 3.1*10<sup>-10</sup> | -0.074 | 0.012 | BMI, smoking initiation, WBC |
| *HTT* | rs362336 | 4:3213832 | G | 0.684 | 0.024 | 0.0037 | 8.3*10<sup>-11</sup> | 0.086 | 0.013 | Depression, WBC, parents survival |
| lncRNA (*LOC105379109*) | rs34073570 | 5:103998895 | C | 0.602 | 0.021 | 0.0035 | 2.1*10<sup>-9</sup> | 0.077 | 0.013 | Autism, depression, loneliness, education, grip strength |
| *HLA-DQB1* | rs28724251 ^ | 6:32629535 | C | 0.649 | -0.041 | 0.0036 | 1.5*10<sup>-30</sup> | -0.156 | 0.013 | ~T2D, ~IBD |
| *HLA-DRB1* | rs139088362 ^ | 6:32539111 | T | 0.961 | -0.068 | 0.012 | 4.3*10<sup>-8</sup> | -0.260 | 0.045 | ~Asthma |
| *HLA-DQA1* | rs36213483 ^ | 6:32611997 | C | 0.761 | -0.049 | 0.005 | 3.4*10<sup>-24</sup> | -0.182 | 0.018 | ~Inflammatory bowel disease |
| *PDE10A* | 6:165102594 | 6:165102594 | TTG | 0.818 | -0.025 | 0.0046 | 4.6*10<sup>-8</sup> | -0.086 | 0.017 | * |
| *FOXP2* | rs2396766 | 7:114318071 | G | 0.527 | -0.022 | 0.0034 | 1.9*10<sup>-10</sup> | -0.078 | 0.012 | "Feeling tense", ~age first birth, ~smoking initiation |
| *HR* | rs56299474 | 8:21992804 | C | 0.827 | -0.025 | 0.0045 | 3.7*10<sup>-8</sup> | -0.087 | 0.016 | * |
| *CSMD3* | rs28453001 | 8:113300930 | G | 0.485 | 0.019 | 0.0034 | 1.7*10<sup>-8</sup> | 0.063 | 0.012 | ** |
| *EXD3* | 9:140251458 | 9:140251458 | G | 0.876 | -0.03 | 0.0051 | 5.5*10<sup>-9</sup> | -0.103 | 0.019 | Education, chronotype |
| *ANK3* | rs4146140 | 10:61885362 | C | 0.617 | 0.019 | 0.0035 | 2.7*10<sup>-8</sup> | 0.067 | 0.013 | ** |
| *NCAM1* | rs10891490 | 11:112885527 | T | 0.405 | 0.02 | 0.0035 | 1.4*10<sup>-8</sup> | 0.066 | 0.013 | Cannabis use, smoking initiation, depression... |
| lncRNA (*LOC105369842*) | rs71460103 | 12:75230067 | G | 0.803 | 0.024 | 0.0043 | 2.3*10<sup>-8</sup> | 0.089 | 0.016 | * |
| *MVK* | rs2111216 | 12:110010151 | A | 0.408 | -0.02 | 0.0035 | 7.7*10<sup>-9</sup> | -0.072 | 0.013 | ~HDL, ~Depression... |
| *INO80* | rs3959554 | 15:41443924 | A | 0.58 | -0.02 | 0.0034 | 9.9*10<sup>-9</sup> | -0.070 | 0.013 | CAD, menopause age, ~SBP |
| *TMOD3* | 15:52168661 | 15:52168661 | CT | 0.536 | 0.019 | 0.0034 | 1.7*10<sup>-8</sup> | 0.074 | 0.012 | ** |
| *PAFAH1B1* | rs7219015 | 17:2555592 | C | 0.782 | -0.023 | 0.0041 | 2.9*10<sup>-8</sup> | -0.084 | 0.015 | * |
| *KRT17P3* | rs3764416 | 17:28900708 | G | 0.129 | -0.029 | 0.0051 | 2.3*10<sup>-8</sup> | -0.101 | 0.019 | * |
| *PHB* | rs10625032 | 17:47472852 | T | 0.754 | 0.022 | 0.004 | 4.4*10<sup>-8</sup> | 0.082 | 0.015 | * |
| *PIK3C3* | rs534731961 | 18:39298720 | G | 0.802 | 0.026 | 0.0043 | 2.5*10<sup>-9</sup> | 0.095 | 0.016 | Smoking initiation, neuroticism |

Abbreviations: RSID = variant identifier; ^ = significantly independent variants at same locus after GCTA analysis; CHR: BP = Genomic position (b37); A1 = effect allele; A1FREQ = A1 allele frequency; BETA = A1 effect on FI (quantile normalized); SE = standard error; P = BOLT LMM p-value; BETA2 = A1 effect on FI (total points, untransformed); SE2 = SE for BETA2.

**Figure 1.**
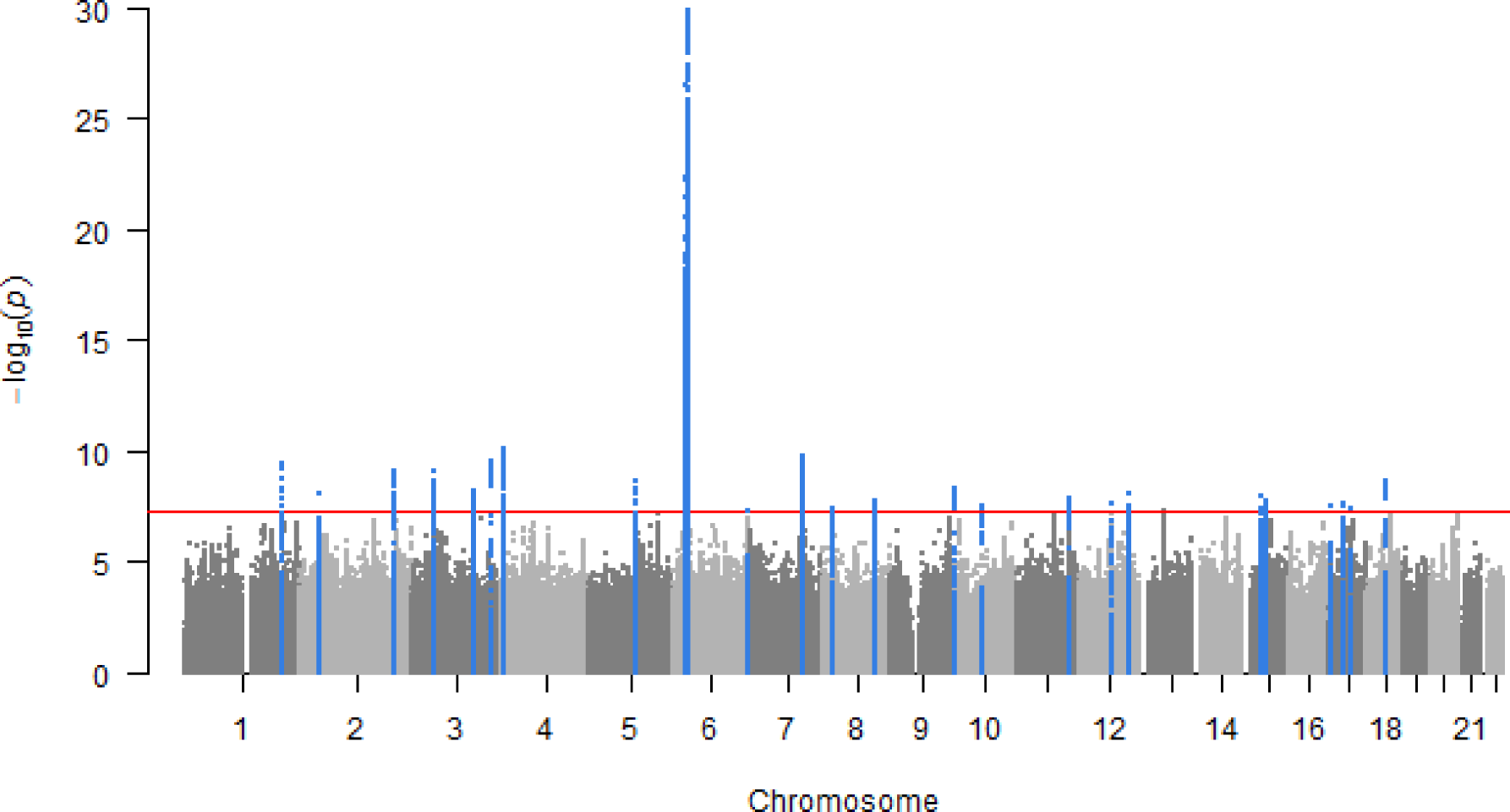
Manhattan plot for genome wide association study of Frailty Index in UK Biobank. GWAS of Frailty Index (normalized) in 164,610 UK Biobank participants aged 60-70 of European descent. Analysis included 16.446.667 autosomal variants with minor allele frequency (MAF) >0.1%. Hardy-Weinberg p-value >1×10^−9^ and imputation quality >0.3. Linear mixed-effects regression models (BOLT-LMM software(Loh et al. 2015). which accounts for relatedness and population structure) were adjusted for age, sex, assessment centre (22 categories) and genotyping array (2 categories: Axiom or BiLEVE). There are 26 independent signals at 24 loci (highlighted in blue) associated with p<5*10^−8^ (red line).

Many of the top 24 loci have previously been associated with traits and diseases in the GWAS Catalog (Buniello et al. 2019) (as of 20th June 2019) such as body mass index (BMI), cardiovascular disease, smoking initiation, HLA proteins, depression, and neuroticism (Table 1, Supplementary Table 1), but nine of the associations appear to be novel. Six of the nine novel loci appear to be less reliable – single variants associated with the FI, with no other correlated variants at the loci showing appreciable evidence of association. Three of the nine novel loci appear more robust and reliable. These are intronic variants in the protein-coding genes *CSMD3* (rs28453001), *ANK3* (rs4146140) and *TMOD3* (single nucleotide deletion on chromosome 15 position 52168661).

### Candidate Gene Association Analysis

Based on prior evidence we also specifically looked up two variants for which we hypothesised an association with the FI *a priori*. Specifically, rs429358, one of the two *APOE* e4 variants – related to both cardiovascular and Alzheimer’s diseases – was not associated with the FI (*p*=0.29). The 9p21.3 variant rs2891168 (associated with parents’ survival in a previous GWAS (RHJ Timmers et al. 2019), but also with heart disease and related traits) in the lncRNA ANRIL – also known as *CDKN2B-AS1* - showed evidence of association, although did not reach genome-wide significance (*p*=3*10^−5^).

### GWAS Replication in TwinGene

We sought replication in 10,616 participants in the TwinGene study. Of the 26 signals from our discovery GWAS, 16 were directly genotyped or imputed in TwinGene, and an additional five had suitable proxy SNPs to use as substitutes (listed in Supplementary Table 2). Two were associated with the FI with consistent directions of estimates: rs759661 and rs7218235 (proxy for rs7219015). However, as power to detect the individual variant associations was limited, we also calculated a weighted genetic risk score (GRS) of the available 21 variants and tested its association with the FI. The GRS was significantly associated with the FI score (β 0.11, 95% CI 0.02-0.20, *p* = 0.015).

### Pathways Enrichment Analysis Implicates Synapse Activity

With gene-set enrichment analysis using MAGMA software, we found four pathways significantly enriched in the GWAS results for FI after adjustment for multiple testing (Bonferroni): “post synapse”, “excitatory synapse”, “synapse”, and “synapse part” (*p*<3*10^−6^; see Supplementary Table 3 for details). In stratified Linkage Disequilibrium Score Regression (LDSR) analysis (Finucane et al. 2018), which identifies whether GWAS statistics are associated with specific gene-tissue pairings in expression or chromatin modification, we found enrichment for the genetic determinants of frailty in several brain regions, including the hippocampus and prefrontal cortex (see Supplementary Tables 4 and 5 for details).

### GWAS hits as molecular quantitative trait loci (QTLs)

We examined whether there are established associations of the top 26 SNPs with concentrations of circulating proteins, gene expression and/or epigenetic markers, as reported by previous QTL GWAS with genome-wide significance (*p*<5*10^−8^). In total, four SNPs are established protein QTLs (Supplementary Table 6). Two *cis*-acting intronic SNPs were associated only with proteins encoded by the genes in which the variants are located (*NCAM1* and *HLA-DQA1*). Two other SNPs – in *HLA-DQB1* and near *RBM6* – were associated in *cis-* or *trans-* with four and five proteins, respectively. 15 of the SNPs were found to be expression QTLs, associated with mRNA varying from one to 14 genes, in up to 43 tissues types (Supplementary Table 7). The most widespread associations with expression were present for rs3764416, which resides in sequence for a non-coding RNA with unknown function on chromosome 17 (*LOC10192709*). In epigenetic marker data (Supplementary Table 8), 16 SNPs were identified as methylation QTLs in one or more blood cell types, with CpG associations per SNP ranging from one to 73. Additionally, four of these SNPs are associated with histone modifications in blood cells, including rs3764416. Variant rs3764416 may also be associated with splicing variation of a nearby pseudogene, *SUZ12P*.

### Methylation - FI associations

Of the total 5,849 variants associated with the FI in UK Biobank, 3,943 were *cis*-methylation quantitative trait loci (*cis*-mQTL) identified in SATSA. These 3,943 variants had 26,851 demonstrable associations with CpG sites in total; single variants were commonly associated with multiple CpG sites, and there were 128 unique CpGs among all associations. Associated variants for the 128 CpGs are shown in Supplementary Table 9. Methylation level in one (cg20614157) of the 128 CpG sites was significantly associated with the SATSA FI after Bonferroni-correction (see Table 2; full results for all 128 sites shown in Supplementary Table 10) and 8 CpG sites were associated with a nominal p<0.05

**Table 2.** Association between methylation levels in mQTL-associated CpGs and Frailty Index in SATSA

| CpG site | Estimate | SE | p | p<sup>Bonferroni</sup> | CHR | BP | UCSC Ref Gene |
| --- | --- | --- | --- | --- | --- | --- | --- |
| cg20614157 | 9.04E-04 | 0.0003 | <0.001 | 0.046 | 6 | 31980845 | *TNXB,TNXA,STK19* |
| cg19376858 | 8.38E-04 | 0.0003 | 0.008 | 1 | 6 | 31980856 | *TNXB,TNXA,STK19* |
| cg15321244 | -2.23E-02 | 0.0088 | 0.011 | 1 | 6 | 32729643 | *HLA-DQB2* |
| cg23928032 | -9.75E-03 | 0.0038 | 0.011 | 1 | 6 | 31964391 | *C4B.C4A* |
| cg01937212 | -1.43E-02 | 0.0062 | 0.020 | 1 | 6 | 32295097 | *C6orf10* |
| cg05139028 | -8.60E-03 | 0.0042 | 0.041 | 1 | 6 | 31323261 | *HLA-B* |
| cg11391305 | -1.43E-02 | 0.007 | 0.042 | 1 | 6 | 32731438 | *HLA-DQB2* |
| cg01248855 | -1.51E-02 | 0.0076 | 0.047 | 1 | 17 | 29234505 | *C17orf42* |

\*If >20 associated variants, the first 20 with lowest p-values are included (see Supplementary Table 10 for full results).
Abbreviations: mQTL=methylation quantitative trait loci; SATSA=Swedish Adoption/Twin Study of Aging; SE=standard error; CHR=chromosome; BP= Genomic position (b37).

### Mendelian Randomization (MR)

To identify potentially modifiable phenotypic traits that may contribute to the development of frailty over the life course, we conducted a series of MR analyses. First, we calculated genetic risk scores (GRS) for 35 traits for which we could retrieve one or more SNPs robustly associated with the traits from pre-existing GWAS results (Figure 2 and Supplementary Table 11). We then tested the association of each GRS with the FI. Using two-sample MR methods, we further examined the top 8 results from GRS analyses that passed Bonferroni correction for 35 tests (p<0.0014): educational attainment, BMI, waist-to-hip ratio, inflammatory bowel disease, grip strength, parental attained age (as a proxy for inherited propensity for longevity), age at menarche and the age at first sexual intercourse (a proxy for a combination of personality characteristics and related health behaviours).

**Figure 2.**
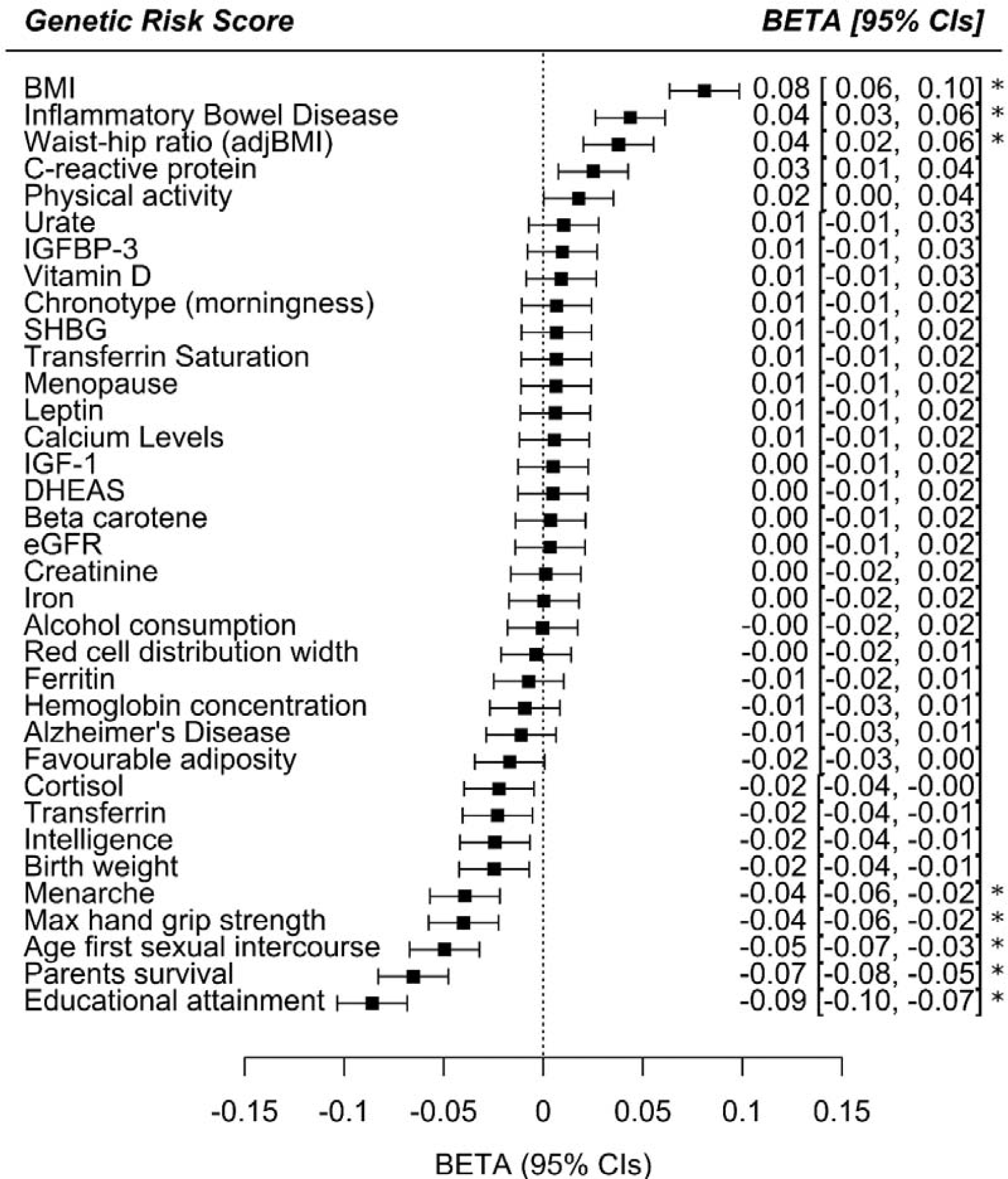
Genetic risk score associations with the frailty index in UK Biobank. Thirty-five exposures, including lifestyle factors, clinical measures, circulating biomarkers and diseases were assessed for their association with the Frailty Index by genetic risk score analysis in UK Biobank participants of European descent aged 60-70 years. Linear regression models included age, sex, assessment centre (22 categories), genotyping array (2 categories: Axiom or BiLEVE) and principal components of ancestry 1-10 as covariates. The betas represent the SD change in FI per SD increase in genetic predisposition to the exposure. Positive betas suggest increased frailty in individuals with greater genetic predisposition to the exposure, whereas negative betas represent a protective effect with increasing genetic predisposition. See Supplementary Table 6 for details. *=significant p<0.0014 after Bonferroni correction for 35 tests. Abbreviations: BMI=body mass index; adjBMI=adjusted for BMI; IGFBP-3=insulin-like growth factor-binding protein 3; SHBG=sex hormone binding globulin; IGF-1=insulin-like growth factor 1; DHEAS=Dehydroepiandrosterone sulfate; eGFR=estimated glomerular filtration rate; CIs=95% confidence intervals.

As trend lines in Figure 3 indicate, educational attainment was estimated to be a determinant of frailty by all model types: on average, a standard deviation increase (i.e. an additional 3.7 years) in education was predicted to lead to 13.6% lower frailty by the seventh decade of life in UK Biobank participants. There was no evidence to suggest that these results were influenced disproportionately by outlying variants, nor biased by directional pleiotropy (MR-Egger intercept test p =0.43), although the test for this may have had limited power. Higher BMI was associated with a higher risk of frailty, and later age of menarche was associated with a lower risk of frailty, although with more modest and inconclusive estimates from MR-Egger and robust/penalised models (see Supplementary Figures 2-8 for scatter plots and full MR results for the non-education phenotypes tests). Model types also produced inconclusive or more divergent findings for grip strength, waist-to-hip ratio, inflammatory bowel disease, parental attained age and age at first sexual intercourse, at least partly reflecting the use of less informative sets of genetic instruments for analyses of several of these traits.

**Figure 3.**
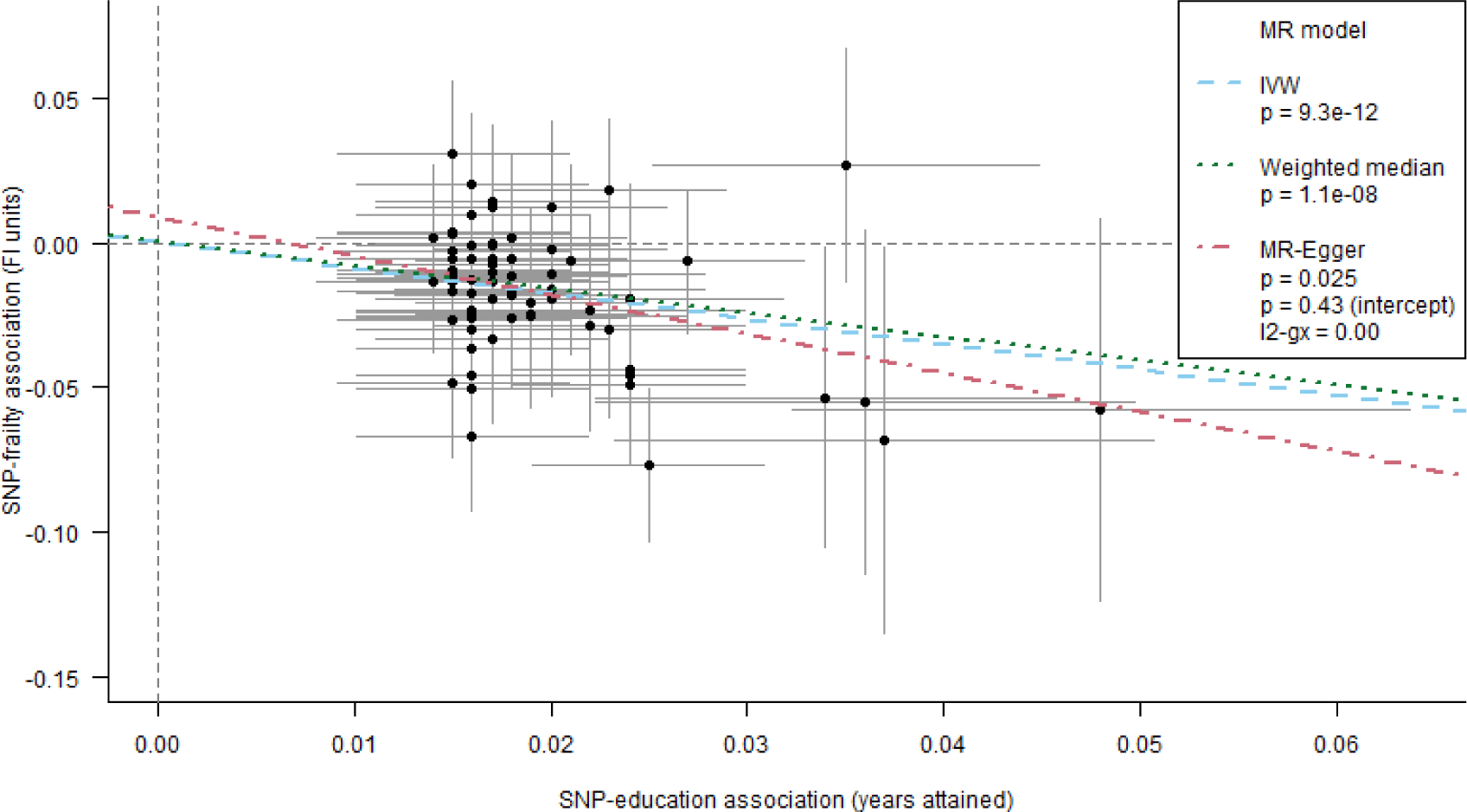
Mendelian randomization estimates for the effect of educational attainment on the frailty index in UK Biobank. Points and error bars represent beta estimates and 95% confidence intervals for each SNP-education / SNP-FI association. The trend lines represent different methods for summarising the estimates from individual SNPS – inverse variance weighting (IVW), weighted median and MR-Egger. The weighted median and MR-Egger estimates are less prone to bias from pleiotropy among the set of variants than IVW, given alternative assumptions hold. The MR-Egger method includes a test of whether the trend’s intercept differs from zero, which indicates whether there is an overall imbalance (directional) of pleiotropic effects: such bias was not identified in this education-FI model.

## DISCUSSION

In this GWAS, we identified 26 independent genetic variants at 24 loci associated with frailty (as measured by the FI) in 164,610 UK community-based individuals aged 60 to 70 years of European ancestry. Many of these loci have previously been associated with traits such as body mass index, cardiovascular disease, smoking initiation, HLA proteins, depression and neuroticism, suggesting roles for known disease risk factors and mental health in the development of frailty, with particular emphasis on synapse maintenance seen in pathways analysis.

Our SNP-based heritability estimate of 14% for the FI was slightly lower than previous family- or twin-based estimates between 19 and 45% (Livshits, Lochlainn, et al. 2018) (Murabito et al. 2012) (Young et al. 2016). This is expected as this SNP-based estimate does not include rare, structural, non-additive, or non-autosomal genetic influences.

Of the 24 loci associated with frailty in this study, most are also associated with traits that have widespread health consequences, such as BMI, smoking initiation, depression, or neuroticism. The results include three independent variants in the major histocompatibility complex region of chromosome six, containing many *HLA* (Human Leukocyte Antigen) genes. HLAs are cell-surface proteins crucial for regulating immune function, which is known to decline with age (Simon et al. 2015). Genetic variants affecting the efficiency of immune function at older ages could have profound consequences for morbidity: variants in this region are robustly implicated in many chronic diseases and traits (Buniello et al. 2019), and also weakness in older people (sarcopenia) (Jones et al. 2019).

In our analysis of blood-based DNA methylation associated with the FI, the only confirmed mQTL CpG (cg20614157) association was in a promotor for the *TNXB/TNXA/STK19* [Tenascin XB/putative Tenascin XA/Serine/threonine-protein kinase 19] gene cluster, located in the *HLA* region. The association of a marker of potential functional relevance with the FI increases the likelihood that one or more of the genes in this cluster are of causal relevance for frailty risk, rather than others in the wider region. We found that high methylation level in cg20614157 was associated with higher frailty; a previous study has found that high methylation of the CpG site is associated with low expression of *TNXB* in breast and skin cancer tissues (Broad Institute of MIT and Harvard 2016). *TNXB* is an extracellular matrix protein thought to be involved in wound healing and collagen function. Further research is needed to examine the potential role of the methylation and expression of *TNXB*, or other genes in this region, with frailty.

To our knowledge, three of the 26 FI-associated variants have not previously been reported in GWAS of any other traits, all in or near genes linked to neuronal function. *CSMD3* (CUB and Sushi multiple domains 3) is expressed in the brain and appears to regulate dendrite development (Mizukami et al. 2016). *ANK3* (Ankyrin 3) is a scaffold protein involved in neurotransmission with links to bipolar disorder and Alzheimer’s disease (Hori et al. 2014) (Franceschi et al. 2018). The final novel locus is *TMOD3* (Tropomodulin 3) which was recently linked to cognitive function (Davies et al. 2018). TMOD3 modulates actin stability, and therefore affects actin binding and remodelling, with diverse downstream effects including cell migration, exocytosis, erythropoiesis, and other developmental pathways (Parreno & Fowler 2018). These results are consistent with FI-defined frailty having a neurological basis, as has been suggested by a previous study showing that frailty and chronic widespread pain have shared neurological pathways (Livshits, Malkin, et al. 2018). Pain questionnaire responses constituted a notable proportion of the FI items in UKB (9 of 49; 18.4%), which will have influenced these findings. However, the relationship of these genetic risk loci with frailty was also present in the replication sample, which included fewer items related to pain perception (2 of 44; 4.5%).

Surprisingly, loci implicated in longevity (RHJ Timmers et al. 2019) or key aging hallmarks (Pilling et al. 2016) were not associated with the FI at genome-wide significance. This included genetic variants in *APOE*, 9p21.3 (*CDKN2A/B*), *TERT*, and *FOXO3A*. The FI is strongly associated with morbidity and mortality epidemiologically, but it may be limited as a tool for identifying specific aging pathways, given the broad definition that includes many diverse diseases and characteristics.

In Mendelian randomization analyses, we tested whether long-term effects of specific physiological, behavioural or lifestyle factors may affect FI scores. The most prominent findings indicated that predispositions to higher educational attainment and lower BMI were related to decreased frailty. Higher BMI directly affects cardiovascular health and subsequent disease risk, in addition to possible indirect effects by reducing socioeconomic status due to stigma (especially in women) (Tyrrell et al. 2016), with subsequent effects on frailty. This also supports a recent study which provided evidence against the notion of an obesity paradox in older people (Bowman et al. 2017).

To our knowledge, this is the largest genetic study of frailty to date. However, this study is limited to participants of European ancestry so results may not be generalizable to other populations. UK Biobank volunteers tend to be healthier and less socioeconomically deprived at baseline than the general UK population (Fry et al. 2017), perhaps reducing power to detect variants associated with the FI. The FI was also based on self-reported baseline data so is open to the possibility of misclassification bias, but the FI has previously been validated in UK Biobank and shown to be strongly predictive of all-cause mortality (Williams et al. 2019). Samples sizes in TwinGene and SATSA are also relatively small so power for the replication and CpG-FI analyses may have been limited.

In conclusion, frailty is influenced by a large number of genetic determinants linked to well-known disease risk factors such as BMI, cardiovascular disease, smoking and HLA proteins. Frailty is also influenced by genetic determinants for depression and neuroticism, suggesting a role for mental health, which may be underpinned by pathways involving synapse maintenance. Future research is required to replicate our results in other cohorts, testing for consistency of genetic determinants when frailty has been measured differently. In particular, a comparable GWAS of the frailty phenotype, based on physical components of health (e.g. exhaustion, weakness) rather than comorbidities, would be likely to yield further risk loci specifically for these components of frailty.

## EXPERIMENTAL PROCEDURES

### UK Biobank

UK Biobank recruited 502,642 community volunteers, aged 37 to 73 years old, from 22 assessment centres across England, Scotland and Wales between 2006 and 2010 (Sudlow et al. 2015). At baseline, participants provided self-reported information on demographics, lifestyle and disease history via questionnaire and underwent physiological measurements, including providing a blood sample for genetics data. We included participants aged 60 to 70 years old at baseline of European descent, with genetics data (*n*=201,062) and available data to generate the frailty index (*n*=164,610) (see Frailty Index section below for details). UK Biobank has ethical approval from the North West Multi-Centre Research Ethics Committee.

### TwinGene

TwinGene data collection took place between 2004 and 2008, when the older participants of the Screening Across the Lifespan Twin (SALT) study were invited to donate blood for molecular and genetic analyses. Both same and opposite-sex twins were included. Ascertainment procedures for SALT (Lichtenstein et al. 2002) and TwinGene (Magnusson et al. 2013) have been previously described. This study was based on the sample that had both genetic and FI data available (*n*=10,616). All participants have given their informed consent. The TwinGene study was approved by the Regional Ethics Review Board, Stockholm.

### Swedish Adoption/Twin Study of Aging (SATSA)

SATSA is a longitudinal study in gerontological genetics, with sampling of participants drawn from the Swedish Twin Registry. SATSA was initiated in 1984 and ended in 2014, and it comprises of nine questionnaire and ten in-person testing (IPT) waves. The participants are same-sex twins, of which some twin pairs have been reared together and some separated before age 11 and reared apart. Ascertainment procedures for SATSA have been described previously (Finkel & Pedersen 2004). This study made use of IPT data, and participants’ first available measurement on FI and DNA methylation were included (*n* =368). All participants have provided informed consent. The SATSA study was approved by the Regional Ethics Review Board in Stockholm.

### Frailty Index (FI)

We used a Frailty Index based on the accumulation of deficits model (Searle et al. 2008), as validated in UK Biobank previously for its ability to predict all-cause mortality (Williams et al. 2019). The FI was derived using 49 self-reported baseline data variables in UK Biobank. Variables were based on a variety of physiological and mental health domains, and included symptoms, disabilities and diagnosed diseases, which were self-reported by participants at baseline (see Supplementary Table 12 for details of the FI components included and the proportion of individuals scoring one for each component). The FI was generated using a complete-case sample with information on all 49 individual components (*n*=164,610) and presented as a proportion of the sum of all deficits. The FI was quantile normalised (i.e. transformed into a normal distribution) prior to the genome-wide association study (due to the skew of the untransformed trait) and sensitivity analyses using the non-transformed trait were performed.

Construction of the FIs for TwinGene and SATSA have been previously described and validated for their ability to predict mortality (Li et al. 2019) (Jiang et al. 2017). Briefly, both the TwinGene FI and SATSA FI were constructed using self-reported questionnaire data (for TwinGene the data collected in SALT were used) and they cover a variety of different health domains. The TwinGene/SALT FI consists of 44 deficits and the SATSA FI consists of 42 deficits. Prior to the analyses, the FIs were quantile-normalised.

### GWAS of the Frailty Index

We used data from UK Biobank v3 genotyping release, described in detail previously (Bycroft et al. 2018). In brief, 488,377 participants were successfully genotyped using custom Affymetrix microarrays for ∼820,000 variants. Imputation was performed using 1000 Genomes and the Haplotype Reference Consortium (HRC) reference panels, yielding ∼93 million variants in 487,442 participants. Participants were excluded if the UK Biobank team flagged them as either having unusually high heterozygosity or missing genotype calls (*n*=968) (Bycroft et al. 2018). Our analysis was restricted to those of European descent (*n*=451,367, method previously published (Thompson et al. 2019)), aged 60 to 70 with complete FI data (*n*=164,610). We analysed 16,446,667 autosomal variants with minor allele frequency (MAF) >0.1%, Hardy-Weinberg p-value >1×10^−9^, and imputation quality >0.3. BOLT-LMM (v2.3.2) software used for the GWAS itself (Loh et al. 2015), which uses linear mixed-effects modelling to account for genetic relatedness and confounding by ancestry. Models included age, sex, assessment centre (22 categories), and genotyping array (two categories: Axiom or BiLEVE) as covariates. We performed conditional analysis to identify independent signals at the same locus using GCTA-COJO (v 1.91.1) (Yang et al. 2012), using a randomly selected subset of 20,000 unrelated UK Biobank participants of European descent as the reference sample. We used Linkage Disequilibrium Score Regression (LDSR, v1.0.0) to estimate the level of bias (e.g. from population stratification and cryptic heritability) in the GWAS, and the heritability of the Frailty Index (Bulik-Sullivan et al. 2015).

### GWAS Replication in TwinGene

The GWAS for the frailty index was replicated in the TwinGene cohort (see Supplementary Methods for details).

### Gene Ontology Pathways & QTL analyses

See Supplementary Methods for details.

### Mendelian Randomization (MR)

MR is the application of genetic variation to infer whether phenotypic traits or exposures affect diseases or health-related outcomes (Lawlor et al. 2008). We investigated a range of exposures for which genetic determinants (typically SNPs) have been identified in previous GWAS. In total, 35 exposure-frailty associations were modelled. Exposures included lifestyle factors, clinical measures, circulating biomarkers and diseases. All traits are listed in Supplementary Table 11, with references for the GWAS from which sets of instrumenting variants were sourced. We used genetic risk scores (GRS) for each trait in initial MR models, and followed up several of the lead findings with sensitivity analyses to detect and account for bias by pleiotropy among genetic instruments (described in the Supplementary Methods).

### Methylation – FI association analysis

To identify examine whether risk variants could be related to frailty via DNA methylation differences, we assessed whether the FI-associated genetic variants harboured methylation quantitative trait loci (mQTL) and whether methylation levels in such loci demonstrated further associations with the FI in SATSA (see Supplementary Methods for further details).

## Data Availability

Data are available on application to the UK Biobank (Sudlow et al. 2015) (www.ukbiobank.ac.uk/register-apply).

## ACKNOWLEDGEMENTS

Access to UK Biobank Resource was granted under Application Number 14631. We would like to thank UK Biobank participants and coordinators for this dataset. We also acknowledge the Swedish Twin Registry for access to data. Genotyping for the Swedish Twin Registry cohorts used in this study was performed by the SNP&SEQ Technology Platform in Uppsala Sweden. The authors would like to acknowledge the use of the University of Exeter High-Performance Computing (HPC) facility in carrying out this work.

## AUTHOR CONTRIBUTIONS

JA derived the FI in UK Biobank and performed descriptive statistics on the data. LP performed GWAS and pathways analysis in UK Biobank. DMW performed QTL look-ups and Mendelian randomization analysis in UK Biobank. JJ performed replication analysis in Twin Gene and mQTL analyses in SATSA. PKM and NLP were responsible of acquisition of the TwinGene genetic data. NLP coordinates the SATSA study, and together with SH, was responsible of acquisition of the SATSA methylation data. YL was responsible for imputing and pre-processing the TwinGene genotype data. YW was responsible of pre-processing the SATSA methylation data. All authors contributed to the design of study, data interpretation and writing of the manuscript.

